# Gut Microbiome Predicts Clinically Important Improvement in Patients with Rheumatoid Arthritis

**DOI:** 10.1101/2020.12.30.20249040

**Authors:** Vinod K. Gupta, Kevin Y. Cunningham, Utpal Bakshi, Benjamin Hur, Harvey Huang, Kenneth J. Warrington, Veena Taneja, Elena Myasoedova, John M. Davis, Jaeyun Sung

## Abstract

**Background:** Rapid advances in the past decade have shown that dysbiosis of the gut microbiome is a key hallmark of rheumatoid arthritis (RA). Yet, the relationship between gut microbiome and clinical improvement in RA disease activity remains unclear. In this study, we explored the gut microbiome of patients with RA to identify features that are associated with, as well as predictive of, minimum clinically important improvement (MCII) in disease activity.

**Methods:** Whole metagenome shotgun sequencing was performed on 64 stool samples, which were collected from 32 patients with RA at two separate time-points. The Clinical Disease Activity Index (CDAI) of each patient was measured at both time-points to assess achievement of MCII; depending on this clinical status, patients were distinguished into two groups. Multiple linear regression models were used to identify microbial taxa and biochemical pathways associated with MCII while controlling for potentially confounding factors. Lastly, a deep-learning neural network was trained upon gut microbiome, clinical, and demographic data at baseline to classify patients according to MCII status, thereby enabling the prediction of whether a patient will achieve MCII at follow-up.

**Results:** We determined that MCII status can explain a significant proportion of the overall compositional variance in the gut microbiome (R^2^ = 3.8%, *P* = 0.005, PERMANOVA). Additionally, by looking at patients’ baseline gut microbiome profiles, we observed significantly different microbiome traits between patients who eventually showed MCII and those who did not. Taxonomic features include alpha- and beta-diversity measures, as well as several microbial taxa, such as *Coprococcus, Bilophila* sp. 4_1_30, and *Ruminococcus* sp. Functional profiling identified thirteen biochemical pathways, most of which were involved in the biosynthesis of L-arginine and L-methionine, to be differentially abundant between the MCII patient groups. In addition to these observations at baseline, we found microbiome features that vary differently in fold-change (from baseline to follow-up) between the two patient groups. These results could suggest that, depending on the clinical course, gut microbiomes not only start at different ecological states, but also are on separate trajectories. Finally, the neural network proved to be highly effective in predicting which patient will achieve MCII (balanced accuracy = 90.0%), demonstrating potential clinical utility of gut microbiome profiles.

**Conclusions:** Our findings confirm the presence of taxonomic and functional signatures of the gut microbiome associated with MCII in RA patients. Ultimately, the gut microbiome may aid in the development of non-invasive tools for predicting future prognosis in RA.

**Trial registration:** N/A

## Background

Rheumatoid arthritis (RA) is a chronic autoimmune inflammatory disease characterized by symmetric polyarticular inflammation and destruction primarily of the synovial joints, as well as of other organ systems [1]. The prognosis of RA has improved over recent decades in parallel with advancements in diagnosis and treatment, particularly the widespread use of biologic and targeted synthetic disease-modifying anti-rheumatic drugs (DMARDs) that enable many persons with RA to achieve low disease activity or clinical remission. However, the exact etiology and pathogenesis of RA are not yet fully understood [2]. In this regard, population-based studies have provided promising evidence that genetic factors contribute to RA onset [3–7]; however, the low concordance rate of RA in monozygotic twins largely suggests the role of non-genetic, environmental factors influencing the incidence of RA [4]. These non-genetic factors include smoking history [8], acute infections [9], and oral and gut microbiota [10].

During the past decade, the role of the gut microbiome in RA pathogenesis has been demonstrated by several experimental studies [11–16]. For example, Maeda *et al*. have shown increased sensitivity to arthritis (via auto-reactive T cell activation in the intestine) in germ-free SKG mice following fecal microbiota transplantation from early RA patients [15]. In addition, another study reported that inflammatory arthritis was strongly attenuated in K/BxN mice under germ-free (GF) conditions; however, the introduction of segmented filamentous bacteria restored splenic auto-antibodies, serum auto-antibodies, and T-helper 17 (Th17) cells [12]. Moreover, the role of gut microbiome in RA pathogenesis is further supported by the attenuation of arthritis in *Il1rn*^−/−^ mice by Tobramycin antibiotic treatment, which led to the decrease in relative abundances of gut commensals, such as *Helicobacter, Flexispira, Clostridium*, and *Dehalobacterium* [16].

Cross-sectional, human gut microbiome studies have elucidated the potential role of gut microbiome ‘dysbiosis’ in RA [13,14,17,18]. A study by Chen *et al*. found lower gut microbial diversity and species richness among RA patients compared to healthy controls; interestingly, patients using methotrexate (MTX) and hydroxychloroquine (HCQ) were observed to have higher gut microbiome diversity and richness than patients not on these medications, possibly indicating partial restoration of normal gut microbiome features with these treatments [13]. Additionally, patients with RA displayed significant improvement in disease activity after being provided with probiotics containing *Bacillus coagulans* [19] or *Lactobacillus casei* [20,21], providing promising evidence towards probiotic therapies in RA treatment. Moreover, another study revealed significant associations between the relative abundance of gut microbial taxa (e.g., Euryarchaeota, Gammaproteobacteria, Erysipelotrichi, and Coriobacteriales) and the disease activity score on 28 joints (DAS28) [22]. Lastly, to demonstrate the potential of targeting the gut microbiome to modulate host immune response and to treat arthritis, Marietta *et al*. have shown that enteral exposure to *Prevotella histicola*, which is a commensal bacterium, in humanized mice can suppress arthritis via mucosal regulation [23].

Certainly, there has been a vast array of recent animal-model studies, cross-sectional case-control studies, and clinical trials showing that a perturbed gut microbiome is a key hallmark of RA. Yet, despite this wide range of novel findings, the association of the gut microbiome with minimum clinically important improvement (MCII) in disease activity in RA patients has yet to be closely examined. The MCII represents the minimal meaningful change (reduction) in quantitative disease activity, and is relevant to patients in terms of improvement in disease symptoms and associated clinical parameters [24]. Although the primary goal in RA management is to achieve and sustain clinical remission or, at least, low disease activity, the MCII in disease activity is also frequently used in clinical settings to evaluate the initial response to treatments. For this, there exists a variety of measurements to quantify RA disease activity, including the Disease Activity Score on 28-joints (DAS28), the Simplified Disease Activity Index (SDAI), and the Clinical Disease Activity Index (CDAI) [25,26]. Among these quantitative indices, the CDAI is one of the most straightforward to use, as it is designed as a simple numerical addition of four components (clinician evaluator global assessment, patient global assessment, 28 swollen joint count, and 28 tender joint count), and does not require acute-phase reactant laboratory tests for its calculation [26].

As medicine evolves towards becoming a big data-centric and bioinformatics-driven discipline [27–29], one of the most promising translational opportunities with gut microbiome datasets arises from their predictive capabilities. In particular, through integrating key biological features (e.g., taxa, functions, genes) of the microbiome with cutting-edge, machine-learning approaches, large-scale data from gut microbiome are positioned to inform various health and wellness applications, and to guide or complement clinical practice. To this point, the gut microbiome has been demonstrated in recent years to facilitate detection of disease [30–34]; classification of disease subtypes and progression stages [35–37]; prediction of clinical outcomes and treatment efficacy [38–42]; personalized nutrition by prediction of postprandial glycemic response [43–45]; and estimation of chronological age [46]. Notably, in a recent study, by applying a random-forest machine-learning model to stool metagenomic data from treatment-naive, new-onset RA patients, Artacho *et al*. found that gut microbiome can aid in the prediction of response to oral administration of methotrexate [47]. Taken together, these examples highlight the potential value of translating microbiome data into new prognostic tools for all areas of precision medicine.

In this study, by investigating the association of gut microbiome profiles from RA patients with MCII and with other patient factors, we demonstrate a computational approach for utilizing gut microbiome information to identify which patients are likely to show clinical improvement independent of baseline clinical features. To this end, we collect shotgun stool metagenomes from a pilot cohort of 32 patients with RA at two separate time-points (i.e., baseline and follow-up). First, we examine the association of gut microbiome with MCII in RA disease activity. Our results show that the status of whether clinical improvement is achieved (or not) is a significant factor contributing to the variance in gut microbiome taxonomic composition. Next, for each time-point, we examine microbiome properties (alpha-and beta-diversity, microbial taxa, and biochemical pathways) that differentiate patients who eventually show clinical improvement from those who do not. Afterwards, we identify taxonomic and functional features whose magnitude and/or direction of change (from baseline to follow-up) varies differently between these two patient groups. Finally, we train a deep-learning neural network model on baseline microbiome, clinical, and demographic data to assess how well we can predict whether MCII in disease activity is attained. Encouragingly, we find that the neural network achieves a 90.0% balanced accuracy in leave-one-out cross-validation, with a compelling accuracy in those who showed clinical improvement (12 correctly predicted among 12 total). Overall, our study offers novel insights into how gut microbial signatures are connected to the trajectory of disease activity in RA, and provides proof-of-concept evidence that accurately forecasting MCII from a stool sample may be possible.

## Methods

### Patient enrollment, eligibility criteria, and sample collection

The study population consisted of consecutive patients with RA attending the outpatient practice of the Division of Rheumatology at Mayo Clinic in Rochester, Minnesota. Eligibility required patients to be adults 18 years of age or older with a clinical diagnosis of RA by a rheumatologist on the basis of the American College of Rheumatology/European League Against Rheumatism 2010 revised classification criteria for RA [48]. Patients were excluded if they did not comprehend English; were unable to provide written informed consent; or were members of a vulnerable population (e.g., incarcerated subjects). On the other hand, patients were eligible irrespective of use of any particular medication. From the patients fulfilling the eligibility criteria, stool samples were collected from two outpatient visits approximately 6–12 months apart, and stored in the ongoing Mayo Clinic Rheumatology Biobank. Clinical and demographic data, including the numbers of tender and swollen joints, patient and evaluator global assessments, C-reactive protein (CRP, mg/L), smoking status, and titers for rheumatoid factor (RF, IU/mL) and anti-cyclic citrullinated peptide antibodies (ACPA), were collected from the electronic medical records. All patients provided written informed consent. The study was approved by the Mayo Clinic Institutional Review Board (no. 14-000616).

### Determination of Minimum Clinically Important Improvement (MCII) in RA disease activity

The CDAI of each patient was measured at two time-points. By taking into account the swollen joint count (of 28 joints), tender joint count (of 28 joints), and the global assessments of disease activity (scored 0–10 on a visual analog scale) by both patient and clinician, the CDAI is scored on a scale ranging from 0 to 76 points [25]. The level of disease activity can be interpreted as low (2.9 ≤ CDAI ≤ 10), moderate (10 < CDAI ≤ 22), or high (22 < CDAI), while CDAI ≤ 2.8 indicates the state of remission [49]. A decrease in CDAI of at least 1 for patients with low disease activity; of at least 6 for patients with moderate disease activity; and of at least 12 for patients with high disease activity between two consecutive visits is considered as MCII in RA disease activity [24]. Based upon this criteria, the study participants can be partitioned into two groups: i) patients who showed clinical improvement (MCII+); and ii) patients who did not show clinical improvement (MCII-).

### Stool sample collection, DNA extraction, and shotgun metagenome sequencing

Stool samples from patients with rheumatoid arthritis were stored in their house-hold freezers (−20°C) prior to shipment on dry ice to the Medical Genome Facility Research Core at Mayo Clinic (Rochester, MN). Once received, the samples were stored at −80°C until DNA extraction. DNA extraction from stool samples was conducted as follows: Aliquots were created from parent stool samples using a tissue punch, and the resulting child samples were then mixed with reagents from the Qiagen Power Fecal Kit. This included adding 60 uL of reagent C1 and the contents of a power bead tube (garnet beads and power bead solution). These were then vigorously vortexed to bring the sample punch into solution and centrifuged at 18000G for 15 min. From there, the samples were added into a mixture of magnetic beads using a JANUS liquid handler. The samples were then run through a Chemagic MSM1 according to the manufacturer’s protocol. After DNA extraction, paired-end libraries were prepared using 500 ng genomic DNA according to the manufacturer’s instructions for the NEBNext Ultra library prep kit (New England BioLabs). The concentration and size distribution of the completed libraries were determined using an Agilent Bioanalyzer DNA 1000 chip (Santa Clara, CA) and Qubit fluorometry (Invitrogen, Carlsbad, CA). Libraries were sequenced at 23–70 million reads per sample following Illumina’s standard protocol using the Illumina cBot and HiSeq 3000/4000 PE Cluster Kit. The flow cells were sequenced as 150 × 2 paired-end reads on an Illumina HiSeq 4000 using the HiSeq 3000/4000 sequencing kit and HiSeq Control Software HD 3.4.0.38. Base-calling was performed using Illumina’s RTA version 2.7.7.

### Quality filtration of sequenced reads

Sequence reads were processed with the KneadData v0.5.1 quality-control pipeline (http://huttenhower.sph.harvard.edu/kneaddata), which uses Trimmomatic v0.36 [50] and Bowtie2 v2.3.2 [51] for removal of low-quality read bases and human reads, respectively. Trimmomatic v0.36 was run with parameters SLIDINGWINDOW:4:30, and Phred quality scores were thresholded at ‘<30’. Illumina adapter sequences were removed, and trimmed non-human reads shorter than 60 bp in nucleotide length were discarded. Potential human contamination was filtered by removing reads that aligned to the human genome (reference genome hg19).

### Taxonomic and functional profiling of stool metagenomes

Taxonomic profiling was performed using the MetaPhlAn2 v2.7.8 [52] phylogenetic clade identification pipeline with default parameters. Briefly, MetaPhlAn2 classifies metagenomic reads to taxonomies based on a database (mpa_v20_m200) of clade-specific marker genes derived from ∼17,000 microbial genomes (corresponding to ∼13,500 bacterial and archaeal, ∼3,500 viral, and ∼110 eukaryotic species). Microbes of viral origin and those that were labeled as either unclassified or unknown were excluded from further analyses. Afterwards, microbiome profiles were normalized using total sum-scaling (TSS) normalization to get the relative abundances (i.e., proportions) of microbial taxonomic ranks.

Functional profiling of annotated MetaCyc biochemical pathways of stool metagenomes was quantified using the HUMAnN v2.8 pipeline [53] with default parameters and with the UniRef90 EC-filtered database integrated into the pipeline. Similarly to the case with taxonomic ranks, MetaCyc pathways unmapped or unintegrated onto the UniRef90 EC-filtered database were discarded from further analyses, and relative abundances of the remaining MetaCyc pathways were calculated using TSS normalization.

### Permutational Multivariate Analysis of Variance based upon taxonomic composition of microbial communities

Bray-Curtis distance matrices based on arcsine, square-root transformed relative abundances of microbial taxa (phylum to species) in stool metagenomes (collected at both clinical visits) were generated using the R ‘vegan’ package v2.5.6. A permutational multivariate analysis of variance (PERMANOVA) [54] was performed on the distance matrix using the ‘adonis2’ function. *P*-values for the test statistic (pseudo-F) were based on 999 permutations to assess the contribution of clinical and demographic characteristics (age group [age < 64 years; age ≥ 64 years], sex [male; female], smoking status [smoker; non-smoker], use of conventional synthetic disease-modifying anti-rheumatic drugs [csDMARDs], use of biologic disease-modifying anti-rheumatic drugs [bDMARDs], use of prednisone, and MCII patient group [MCII+; MCII-]) to the total variance in gut microbial community composition. (of note, categorical age group was used due to the uneven and skewed distribution of continuous age.) Intra-subject longitudinal variation was accounted for by constraining permutations to within visits using the ‘strata’ argument. Both marginal (i.e., univariate analysis) and adjusted (i.e., multivariate analysis controlling for multiple covariates simultaneously) models were used to evaluate percent variance and significance of associations between gut microbiome composition and patient factors.

### Comparisons of alpha- and beta-diversity between MCII patient groups

Overall ecology of gut microbiomes was evaluated by calculating alpha-diversity (Fisher’s Index and richness) and beta-diversity (Bray-Curtis distance between all sample-pairs) based upon untransformed relative abundances of microbial species in each stool metagenome using the R ‘vegan’ package v2.5.6. Multiple linear regression models (MLRMs) were then constructed using the R ‘stats’ package v3.6.3 to determine the alpha-diversity indices that were significantly different between MCII+ and MCII-groups. MLRMs were adjusted for clinical and demographic characteristics that explained significant (or nearly significant) proportions of the variance in gut microbial community composition. Mann-Whitney *U* test was used to evaluate the statistical significance of the difference in beta-diversity between the patient groups.

### Identification of differentially abundant microbial taxa and biochemical pathways between MCII patient groups

To identify differentially abundant microbial taxa and biochemical pathways between MCII+ and MCII-groups (at either baseline or follow-up), MLRMs were constructed for arcsine, square-root transformed relative abundance of each taxon and pathway. Only taxa and pathways that were detected in at least a third of all samples were included for analysis (resulting in a total of 176 and of 262, respectively). All MLRMs were designed to model the relationship between a taxon/pathway and MCII patient group, while adjusting for clinical and demographic characteristics found to be significantly (or nearly significantly) associated with gut microbiome compositional variance according to the aforementioned PERMANOVA analysis. Taxa and pathways were considered as differentially abundant between the two MCII patient groups if the corresponding regression coefficient for the patient group was significant (*P* < 0.05).

### Quantification of fold-change in gut microbial taxa and biochemical pathways

Microbial taxa and biochemical pathways detected in at least a third of all samples were considered for the calculation of fold-change (log_2_(FC)) from baseline to follow-up visit. As log_2_(FC) cannot be applied to cases with zeros (which are often abundant in microbiome relative abundance data), a pseudo-count (1.0 × 10^−5^) was added to all relative abundances, as suggested by Kaul *et al*. [55]. Then, MLRMs were designed for each taxon and pathway to identify any significant differences (*P* < 0.05) in log_2_(FC) of relative abundances between the two MCII patient groups. All MLRMs were adjusted for clinical and demographic characteristics found to be significantly (or nearly significantly) associated with gut microbiome compositional variance according to the aforementioned PERMANOVA analysis.

### Construction of neural networks for predicting MCII and CDAI

Two separate multi-layer (deep) feedforward artificial neural networks with stochastic gradient descent using back-propagation, which were provided by the Python version of the ‘H2O’ package v3.26.0.3, were constructed to meet the following two objectives (i.e., output layer): i) classify a patient as MCII+ or MCII-from all baseline gut microbiome (relative abundances of 176 taxonomic ranks and of 262 MetaCyc pathways), clinical (CDAI, use of medications [bDMARDs, csDMARDs, and prednisone], HAQ, pain, and CRP), and demographic data (age, sex, and smoking status). In other words, predict whether a patient will achieve MCII based on all identifiable baseline features. This model’s predictive performance was evaluated by leave-one-out cross-validation on all baseline profiles; and ii) predict CDAI using the aforementioned microbiome, clinical (except for CDAI), and demographic data as input predictor variables for the neural network. Predictive performance of this model was evaluated by a leave-one-patient-out cross-validation method. More specifically, in each cross-validation loop, both samples from the same patient were allocated as an internal validation set, while all remaining samples were used as the internal training set for constructing the neural network to predict CDAI scores of the allocated two samples. For both objectives, the default input parameters were used for model-training except for the following: Epochs = ‘10,000’; and Random seed = ‘1234’. See http://docs.h2o.ai/h2o/latest-stable/h2o-docs/data-science/deep-learning.html for all parameters of the neural network and their default values. Data curation and model implementation was performed in Python v3.6.4 on individual cloud instances utilizing Amazon Web Services (AWS).

## Results

### Study population

From a total of 86 patients with RA whose blood and/or stool samples were stored in our ongoing biobank, we identified 51 patients who had at least two available stool samples collected at least 6 to 12 months apart (102 total samples). From these 51 patients, we found 36 patients (72 samples) who had fully available clinical data (to assess CDAI and MCII) and demographic information at both clinical visits, thereby leading to the exclusion of 15 patients (30 samples). We excluded an additional 4 patients (8 samples) from further analysis because they were in clinical remission at both clinical visits. Hence, this retrospective, observational cohort study includes 32 participants (64 samples), of whom 65.6% (21 of 32) were female.

At the time of baseline stool sample collection, the patients had established disease with a mean age of 64.9 years (s.d. = 11.0), and a mean disease duration of 8.2 years (s.d. = 8.2). A summary of the patient enrollment, eligibility criteria, and sample collection protocol is provided in **Methods**. At baseline, all patients were on treatment with biologic disease-modifying anti-rheumatic drugs (bDMARDs, 46.9%), conventional synthetic disease-modifying anti-rheumatic drugs (csDMARDs, 87.5%), or prednisone (46.9%). For any medication, no association was found between its use at baseline and MCII in RA disease activity (**Fig. S1**). Baseline and follow-up visits were separated by a mean duration of 9.5 months (s.d. = 3.6 months), which was numerically longer for patients who attained MCII than for patients who did not attain MCII though not statistically significant (median 363 vs. 252 days, respectively; *P* = 0.08, Mann-Whitney *U* test). At all instances of stool sample collection, disease activity of patients varied from remission to high disease activity, with a mean CDAI of 16.3 (s.d. = 13.7) and 13.6 (s.d. = 11.6) at baseline and follow-up, respectively.

**Figure S1.**
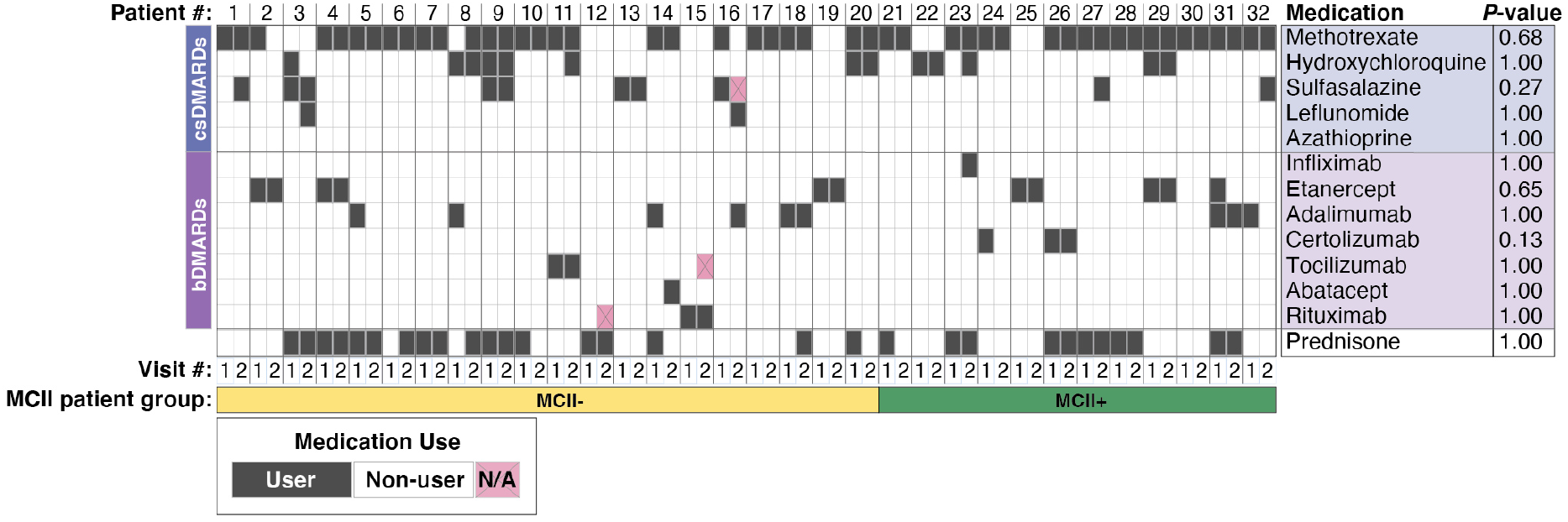
Overview of medication use by all 32 study participants. *P*-values from Fisher’s exact test (right) indicate statistical significance of association between MCII patient group and baseline medication use/non-use. No significant association for any medication was found. bDMARDs, biologic disease-modifying anti-rheumatic drugs. csDMARDs, conventional synthetic disease-modifying anti-rheumatic drugs. MCII, minimum clinically important improvement. MCII+, patients who showed MCII. MCII-, patients who did not show MCII.

In total, 12 of the 32 (37.5%) total study participants achieved MCII in RA disease activity at their follow-up visit. The average change in CDAI for these 12 patients was −16.7 (s.d. = 12.8) units, which was, as expected, significantly different from the average change in CDAI of 5.7 (s.d. = 8.9) units for the remaining 20 of 32 (62.5%) patients who did not show improvement in RA disease activity (*P* = 6.9 × 10^−6^, Mann-Whitney *U* test). We used Fisher’s exact test to identify significant differences in categorical variables (e.g., age group, sex, smoking status, medication use, presence of rheumatoid factor and of anti-cyclic citrullinated peptides antibodies), and Mann-Whitney *U* test to identify significant differences in continuous clinical measurements (CDAI, health assessment questionnaire [HAQ], swollen joint count [SJC], tender joint count [TJC], C-reactive protein [CRP], patient’s and physician’s health status assessment) between two patient groups: MCII+ (i.e., patients who showed MCII in disease activity based upon the change in CDAI from baseline to follow-up visit) and MCII-(i.e., patients who did not show MCII) (**Table 1**). At baseline, we found significant or nearly significant associations between MCII patient group (i.e., MCII+ and MCII-) and CDAI (*P* = 0.03), provider global evaluation of disease activity (md_vas) (*P* = 0.06), and CRP (*P* = 0.06). At follow-up visit, we found the following factors to be significantly associated with MCII patient group: CDAI (*P* = 1.9 × 10^−3^), change in CDAI from baseline (*P* = 6.9 × 10^−6^), pain (VAS) (*P* = 2.8 × 10^−3^), TJC (*P* = 0.01), patient global evaluation of disease activity (pt_vas) (*P* = 6.3 × 10^−3^), and provider global evaluation of disease activity (md_vas) (*P* = 0.01).

**Table 1.**
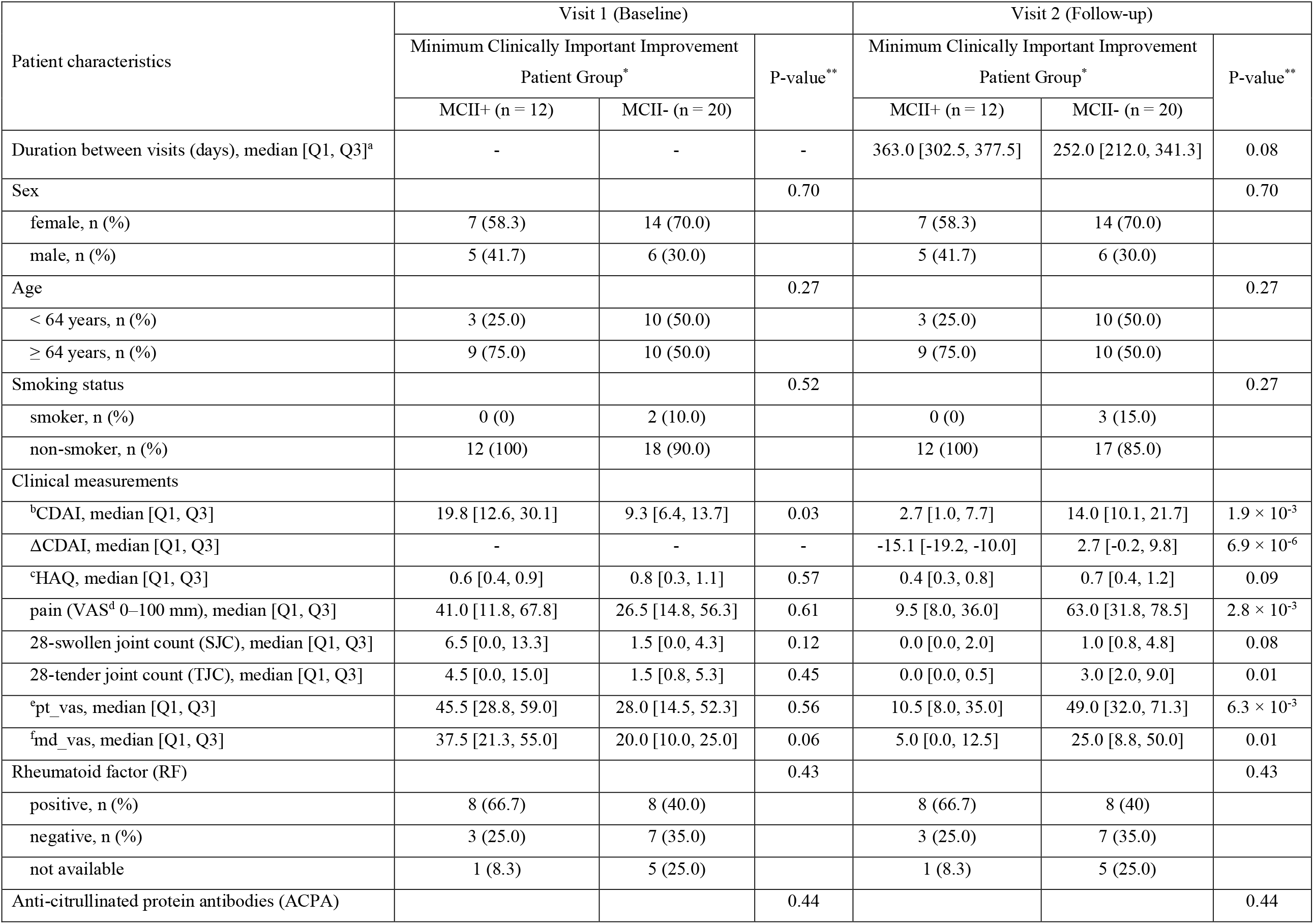
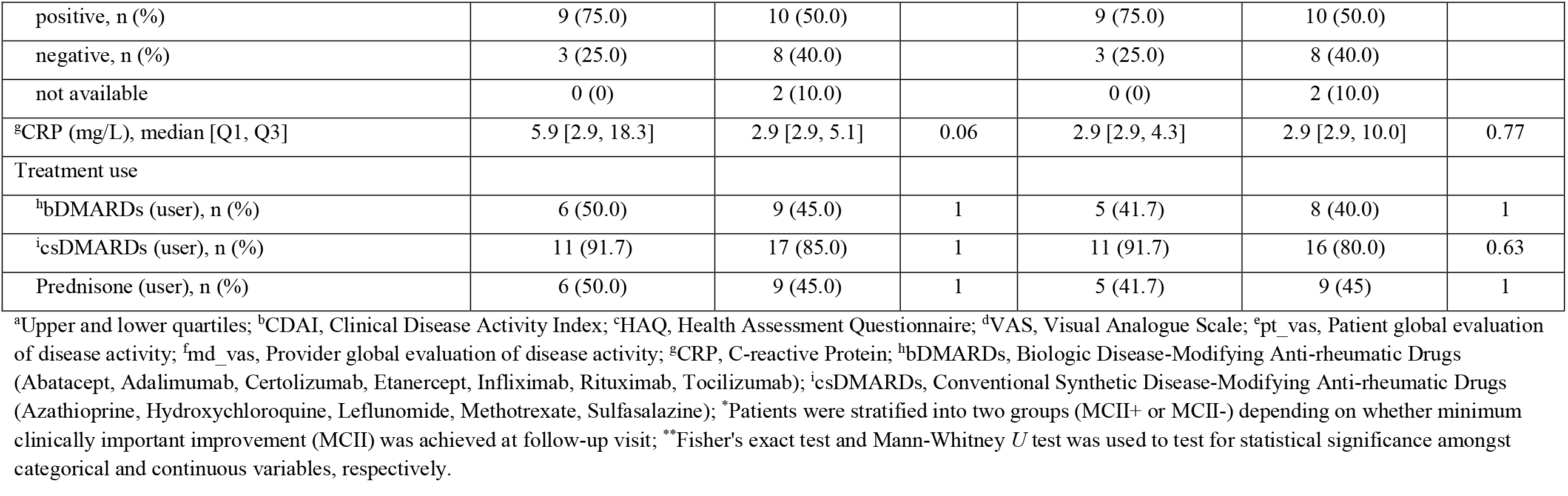
Demographic and clinical characteristics of the study population.

### MCII patient group explains significant variance in gut microbial community composition

We performed a PERMANOVA analysis to evaluate the patient characteristics that contribute to the variance in gut microbial communities of patients with RA (**Methods**). Using univariate (marginal) models, as well as multivariate (adjusted) models that jointly take into consideration all measurable factors, we considered MCII patient group, age group, sex, smoking status, baseline CDAI, and medication use (for csDMARDs, bDMARDs, and prednisone). Of note, we assume that the resulting percent variance explained by each variable in the adjusted model is statistically independent of other variables.

We found that MCII patient group explained 3.8% of the total variance in gut microbial communities (*P* = 0.005, PERMANOVA; **Table 2** and **Fig. 1a**), after controlling for age group, sex, smoking status, CDAI, use of csDMARDs (all of which explained significant or nearly significant variance in marginal models), and intra-subject longitudinal variation. The adjusted model also showed that age group, use of csDMARDs, sex, smoking status, and CDAI explained 7.7%, 3.1%, 2.9%, 2.7%, and 2.3% of the total variance, respectively (**Table 2** and **Figs. 1b–f**), indicating partial dependence of gut microbiome profiles among patients with RA on these other factors. However, treatment with bDMARDs (*P* = 0.23, PERMANOVA; **Fig. 1g**) or prednisone (*P* = 0.26, PERMANOVA; **Fig. 1h**) was not found to have any significant association with gut microbial community composition (**Table 2**). Taking into account these observations, we controlled for age group, use of csDMARDs, sex, smoking status, and CDAI in subsequent analyses for investigating the differences in gut microbiome profiles between patients of the MCII+ group and those of the MCII-group.

**Table 2.** Patient characteristics contributing to the variance in gut microbial community composition.

| Patient characteristics<sup>¶</sup> | Marginal model |  | Adjusted model |  |
| --- | --- | --- | --- | --- |
| | Variance (%) | $P$-value<sup>#</sup> | Variance (%) | $P$-value<sup>#</sup> |
| Age group | 7.7 | 0.001 | 7.7 | 0.001 |
| MCII patient group | 4.4 | 0.004 | 3.8 | 0.005 |
| csDMARDs | 3.7 | 0.010 | 3.1 | 0.010 |
| Sex | 3.1 | 0.024 | 2.9 | 0.014 |
| Smoking status | 4.0 | 0.003 | 2.7 | 0.023 |
| CDAI | 2.3 | 0.130 | 2.3 | 0.061 |
| bDMARDs | 1.8 | 0.278 | 1.6 | 0.228 |
| Prednisone | 1.7 | 0.332 | 1.6 | 0.260 |

<sup>¶</sup>Each patient characteristic was measured for 32 patients at both clinical visits. All 64 gut microbiome samples were analyzed simultaneously using Permutational Multivariate Analysis of Variance (PERMANOVA); <sup>#</sup>PERMANOVA was used to test for statistical association between corresponding patient characteristic and variance within microbiome composition.

**Figure 1.**
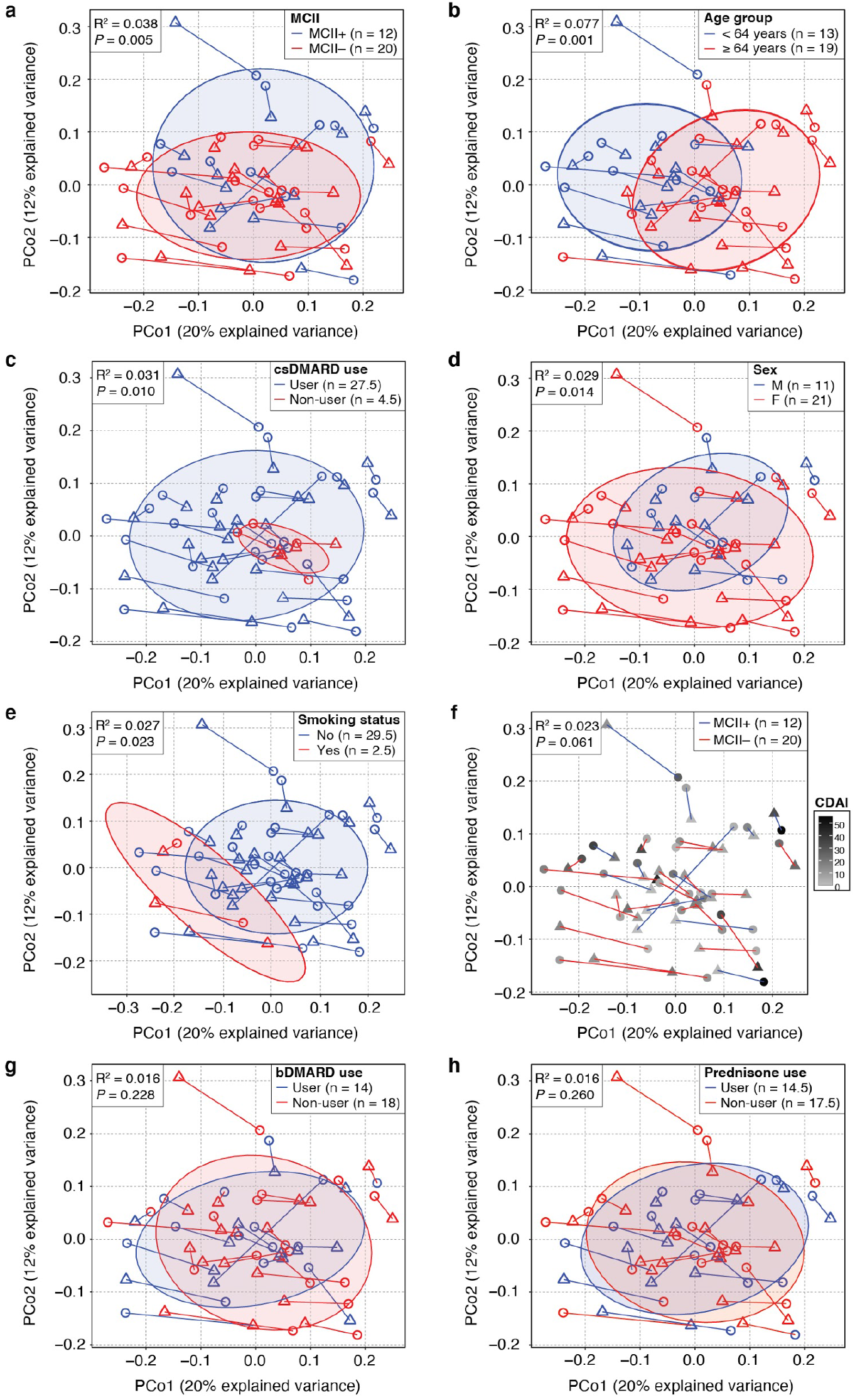
Principal Coordinates Analysis (PCoA) ordination plots of gut microbiome samples from patients with RA (*n* = 32). PERMANOVA analysis finds that the variance in gut microbial community composition can be explained by **(a)** MCII patient group, **(b)** age group, **(c)** use of csDMARDs, **(d)** sex, **(e)** smoking status, and **(f)** CDAI, but by neither **(g)** use of bDMARDs nor **(h)** use of prednisone. All 64 gut microbiome samples (from 32 patients at both clinical visits) were analyzed simultaneously using PERMANOVA, while intra-subject longitudinal variation was accounted for by constraining permutations to within visits. R^2^ and *P*-values derived from PERMANOVA. Each circle and triangle signifies baseline and follow-up, respectively. Lines connect time-points of the same patients. Ellipses correspond to 80% confidence regions. MCII, minimum clinically important improvement. MCII+, patients who showed MCII. MCII-, patients who did not show MCII. Non-integer ‘n’ corresponds to cases wherein the patient reported differently at baseline than at follow-up.

### Gut microbial taxa show significant associations with clinical disease activity

We investigated the association between the relative abundance of gut microbial taxa and quantitative disease activity (CDAI), while controlling for the aforementioned covariates. We found that thirteen taxa, including Erysipelotrichia (class); Bacteroidaceae (family); *Anaerotruncus* and *Bacteroides* (genus); *Anaerotruncus colihominis, Clostridium spiroforme*, and *Pseudomonas mendocina* (species), were significantly associated with CDAI (**Fig. S2**). Among these thirteen, ten microbial taxa were found to be positively correlated with CDAI, whereas three (Bacteroidaceae, *Bacteroides*, and *Lachnospiraceae bacterium* 3_1_46FAA) displayed negative correlations.

**Figure S2.**
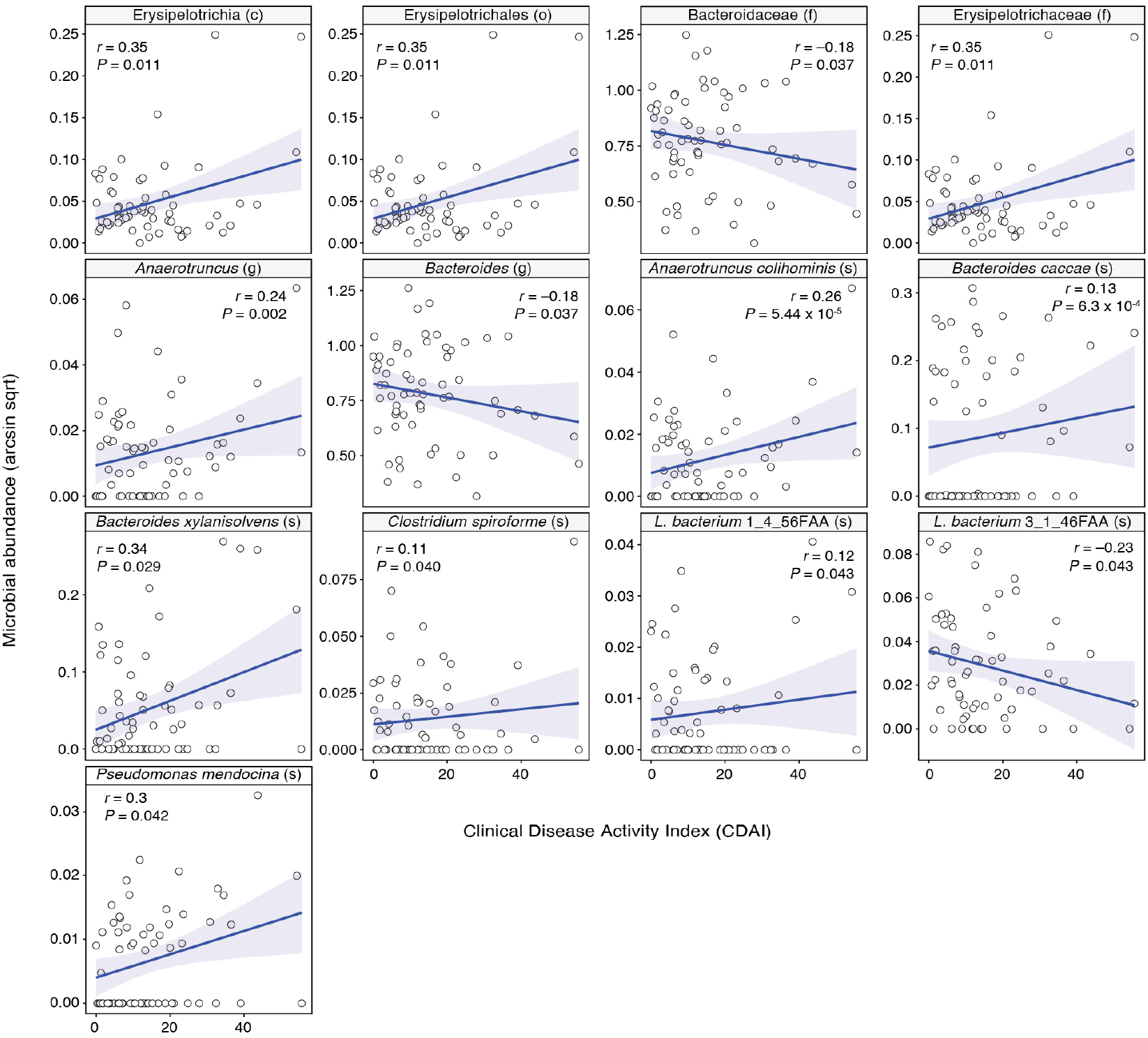
Microbial taxa having significant associations with Clinical Disease Activity Index (CDAI). Points within the scatter-plots represent gut microbiome samples (64 total samples from 32 patients at both clinical visits), while blue lines indicate the best fit. *r* corresponds to the Pearson correlation coefficient between CDAI and arcsine, square-root transformed microbial taxa relative abundance. *P*-value, which shows the significance of the association between CDAI and taxa abundance, corresponds to the regression model coefficient for CDAI in a mixed-effects linear regression model adjusting for fixed effects (age group, sex, smoking status, use of csDMARDs) and random effects (Patient ID). Taxonomic ranks: c, class; o, order; f, family; g, genus; s, species.

### Features of baseline gut microbiomes significantly differ between MCII+ and MCII-patient groups

At baseline, we observed Bacteroidetes and Firmicutes as the most abundant phyla based upon relative abundances (**Fig. 2a**); Bacteroidales and Clostridialis as the most abundant orders (**Fig. 2b**); and Bacteroidaceae as the most abundant family (**Fig. 2c**). We next investigated the baseline gut microbiomes of all 32 patients to identify differences in ecological properties (e.g., alpha-/beta-diversity) or in individual taxonomic and functional features between the two MCII patient groups. In effect, by knowing—albeit retrospectively—the clinical outcomes in advance, we have asked: on the basis of gut microbiome information, can differences at baseline not only provide hypotheses that connect gut microbiome to clinical improvement, but also reveal biomarkers predictive of the clinical course? To this end, we found higher species-level alpha-diversity, that is, Fisher’s Index (*P* = 0.002, MLRM; and **Fig. 3a**) and richness (*P* = 0.0004, MLRM; and **Fig. 3b**), and higher beta-diversity, that is, Bray-Curtis distances between all pairs of samples (*P* = 0.002, Mann-Whitney *U* test; and **Fig. 3c**) in the MCII+ group compared to the MCII-group. In addition, we sought to identify microbial taxa and microbiome-derived annotated MetaCyc biochemical pathways that were differentially abundant between the two MCII patient groups at baseline. Our analysis led to the following ten microbial taxa, all of which were found to be higher in the MCII+ group: Negativicutes (class), Selenomonadales (order), Prevotellaceae (family), *Coprococcus* (genus), *Bacteroides* sp. 3_1_19 (species), *Bilophila* sp. 4_1_30 (species), *Blautia* sp. KLE_1732 (species), *Coprococcus comes* (species), *Ruminococcus* sp. (species), and *Streptococcus salivarius* (species) (*P* < 0.05, MLRM; and **Fig. 3d**). Moreover, we found thirteen MetaCyc pathways that were differentially abundant between MCII+ and MCII-groups at baseline (*P* < 0.05, MLRM; and **Fig. 3e**). Six of these pathways, which include multiple for tetrahydrofolate biosynthesis and L-methionine biosynthesis, were significantly higher in patients of the MCII+ group than in those of the MCII-group; in contrast, the remaining seven pathways, the majority of which being for L-arginine and L-ornithine biosynthesis, were more abundant in patients of the MCII-group. Taken together, our results show that gut microbiomes of the two diverging patient groups start at different ecological states even before reaching their clinical endpoints.

**Figure 2.**
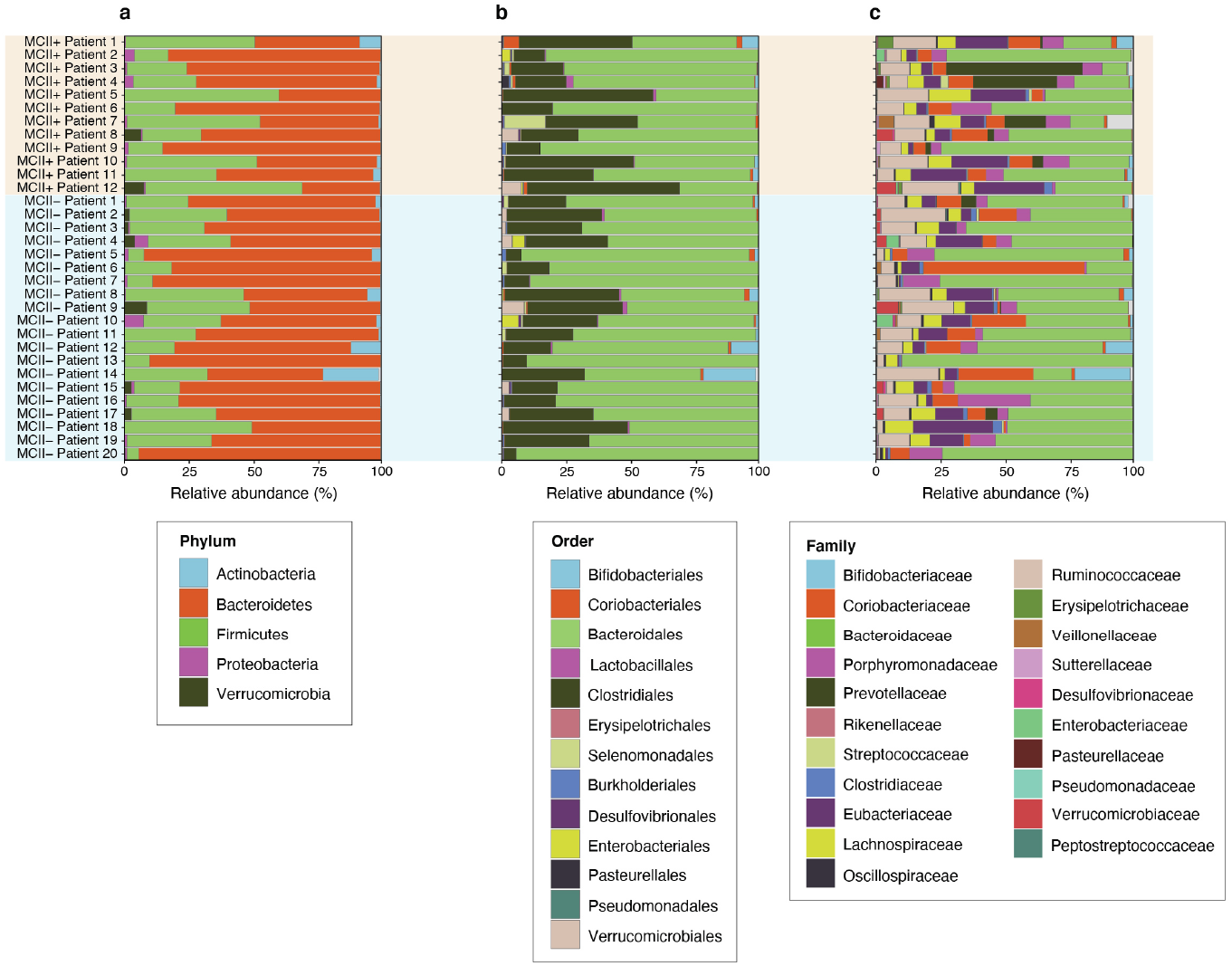
Stacked bar-plots showing the distribution of relative abundances of taxonomic ranks detected in baseline gut microbiomes. At **(a)** phylum-level, Bacteroidetes and Firmicutes were the two most abundant phyla. At **(b)** order-level, Bacteroidales and Clostridiales were most abundant. Among **(c)** families, Bacteroidaceae was the most abundant.

**Figure 3.**
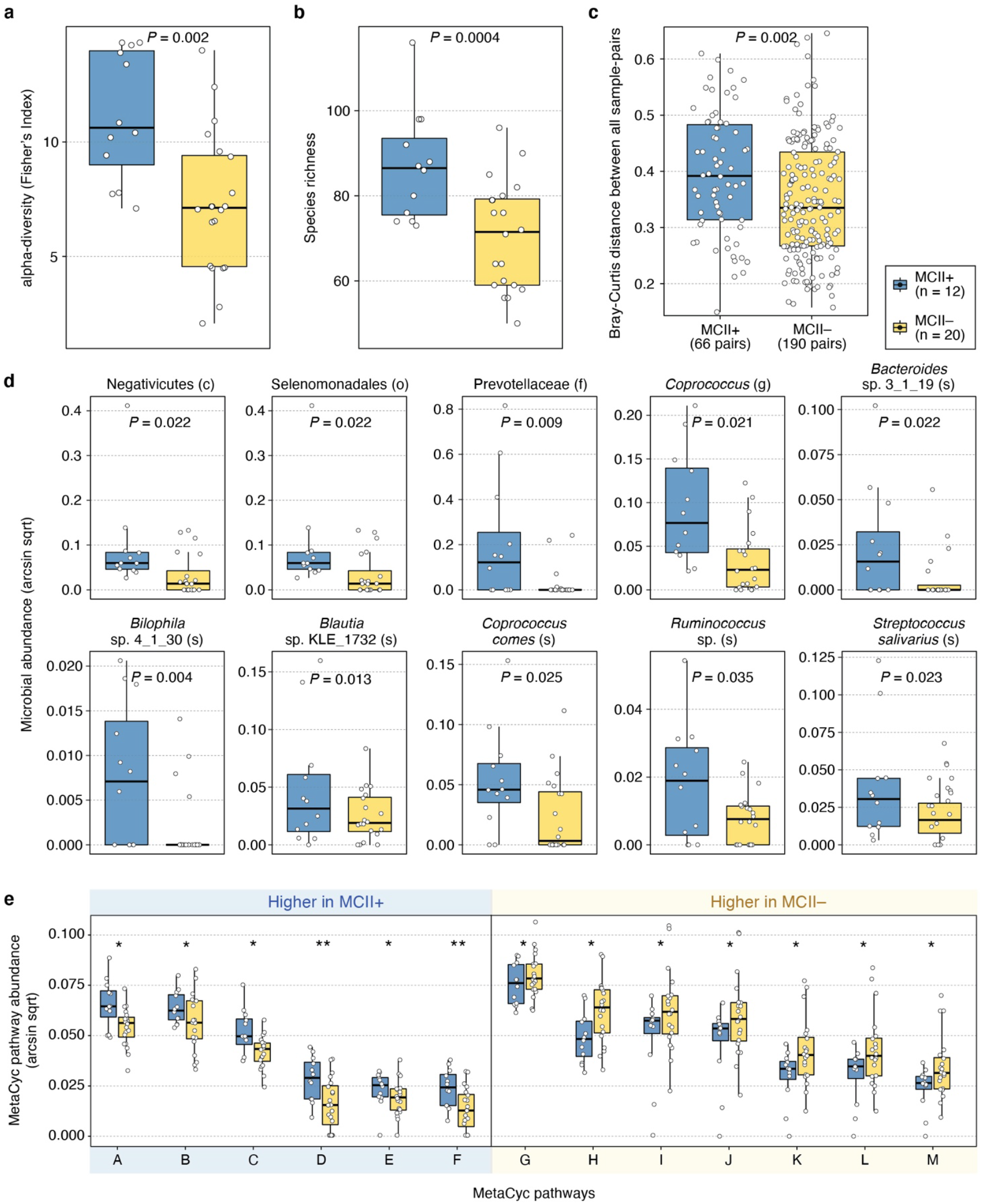
Differences in baseline gut microbiome features between MCII+ and MCII-patient groups. For alpha-diversity, significantly higher species-level **(a)** Fisher’s Index (*P* = 0.002, MLRM) and **(b)** richness (*P* = 0.0004, MLRM) were observed in the MCII+ group. **(c)** In regard to beta-diversity, a higher distribution of Bray-Curtis distances between all sample-pairs was found in the MCII+ group (*P* = 0.002, Mann-Whitney *U* test). **(d)** Ten microbial taxa were identified as differentially abundant between the two patient groups (*P* < 0.05, MLRM). All ten were significantly higher in abundance in the MCII+ group. **(e)** Thirteen MetaCyc biochemical pathways were found to be differentially abundant. Except for beta-diversity, multiple linear regression models (MLRMs) were designed to test for the statistical significance of the relationship between MCII patient group and microbiome features, while controlling for patient factors (age group, sex, smoking status, CDAI, and use of csDMARDs) reported at baseline. *P*-value corresponds to the regression model coefficient for MCII patient group. ^*^, 0.01 ≤ *P* < 0.05; ^**^, 0.005 ≤ *P* < 0.01. MCII, minimum clinically important improvement. MCII+, patients who showed MCII. MCII-, patients who did not show MCII. Taxonomic ranks: c, class; o, order; f, family; g, genus; s, species. MetaCyc pathways: A, L-homoserine and L-methionine Biosynthesis; B, L-tryptophan Biosynthesis; C, L-methionine Biosynthesis I; D, Superpathway of Tetrahydrofolate Biosynthesis and Salvage; E, Formaldehyde Assimilation III; F, Superpathway of Tetrahydrofolate Biosynthesis; G, Adenine and Adenosine Salvage III; H, CMP-3-deoxy-D-manno-octulosonate Biosynthesis I; I, L-arginine Biosynthesis IV; J, L-arginine Biosynthesis I; K, L-arginine Biosynthesis III; L, L-arginine Biosynthesis II; M, L-ornithine Biosynthesis.

In contrast to the case at baseline, we observed no significant differences in species-level Fisher’s Index (*P* = 0.07, MLRM), richness (*P* = 0.20, MLRM), and Bray-Curtis distances between all sample-pairs (*P* = 0.31, Mann-Whitney *U* test) between the two MCII patient groups at follow-up visit (**Figs. S3a–c**). However, we found eight taxa to be differentially abundant: Negativicutes (class), Selenomonadales (order), Prevotellaceae and Veillonellaceae (family), and *Veillonella* (genus) were higher in the MCII+ group; and *Bacteroides uniformis, Clostridium leptum*, and *Erysipelotrichaceae bacterium* 6_1_45 (species) were higher in the MCII-group (*P* < 0.05, MLRM; **Fig. S3d**). Furthermore, we identified seven differentially abundant biochemical pathways (*P* < 0.05, MLRM; **Fig. S3e**), among which those for glycogen degradation, putrescine biosynthesis, polyamine biosynthesis, sulfate assimilation, and cysteine biosynthesis were higher in the MCII+ group.

**Figure S3.**
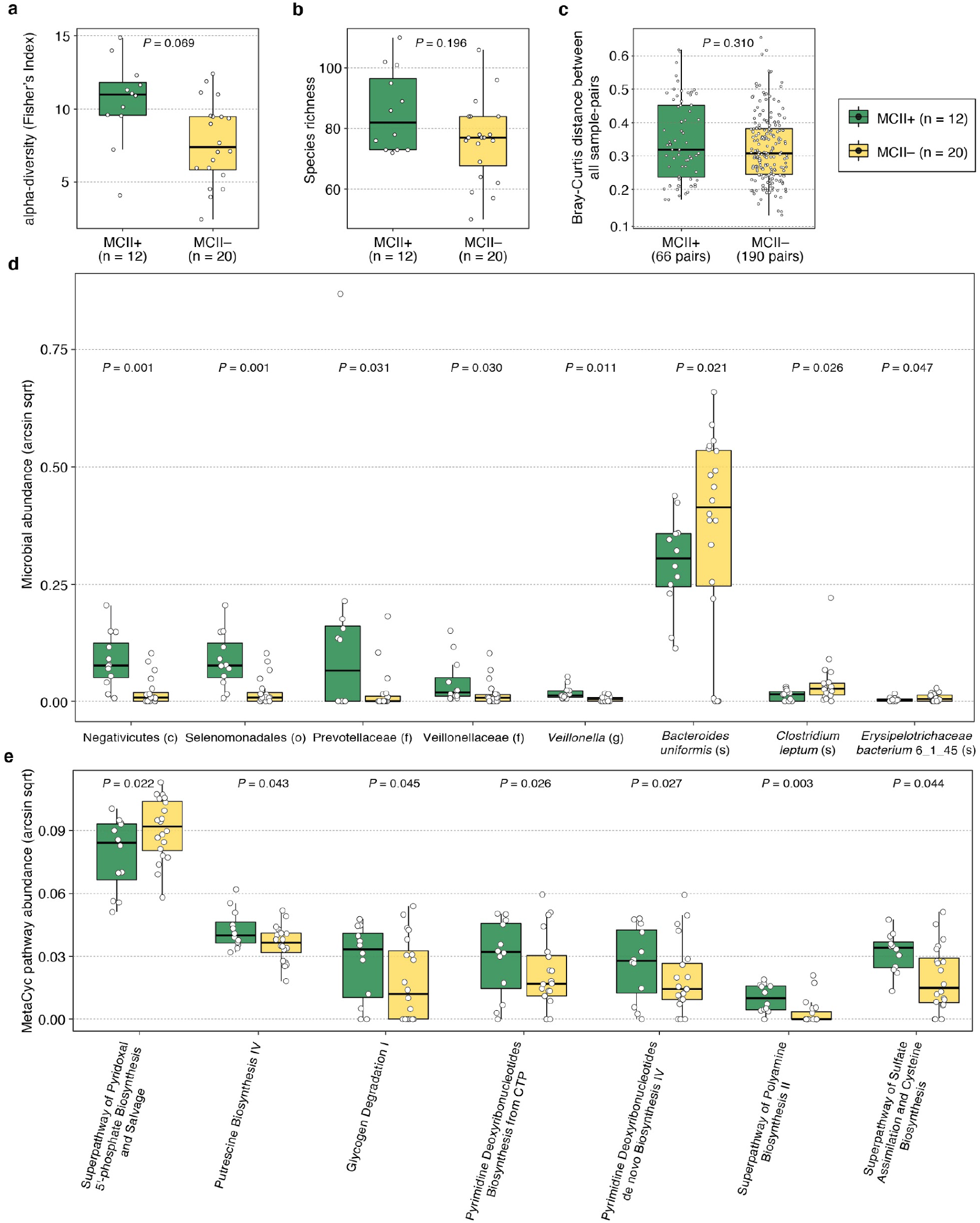
Differences in gut microbiome features between MCII patient groups at follow-up visit. In the gut microbiomes of patients with RA at follow-up, species-level **(a)** Fisher’s Index (*P* = 0.07, MLRM), **(b)** richness (*P* = 0.20, MLRM), and **(c)** Bray-Curtis distances between all sample-pairs (*P* = 0.31, Mann-Whitney *U* test) were not found to be significantly different between the MCII+ and MCII-patient groups. **(d)** Eight microbial taxa and **(e)** seven MetaCyc biochemical pathways were identified as differentially abundant between the two MCII groups while controlling for patient factors (age group, sex, smoking status, CDAI, and use of csDMARDs) reported at follow-up. Microbial taxa and biochemical pathways were considered as differentially abundant if the regression model coefficient for MCII patient group was significant (*P* < 0.05). MCII, minimum clinically important improvement. MCII+, patients who showed MCII. MCII-, patients who did not show MCII. Taxonomic ranks: c, class; o, order; f, family; g, genus; s, species.

### Gut microbiome taxa and functions show significant differences in fold-change from baseline to follow-up between MCII patient groups

We examined the longitudinal variation in relative abundances (i.e., fold-change from baseline to follow-up) of microbial taxa and of biochemical pathways. From this, we sought to identify differences in how the gut microbiome changes in association with clinical outcomes (i.e., showing clinical improvement or not). First, we found that patients of the MCII+ and MCII-groups showed significant fold-change differences in the following six microbial taxa (*P* < 0.05, MLRM; **Fig. 4a, Fig. S4a**): i) fold-change in *Oscillibacter* (genus) was higher in the MCII+ group. This result suggests that *Oscillibacter* increases in relative abundance more highly and/or frequently in the MCII+ group compared to the MCII-group; and ii) fold-changes in *Coprococcus* (genus), *Ruminococcus* (genus), *Clostridium leptum* (species), *Oscillibacter* sp. KLE_1728 (species), and *Streptococcus thermophilus* (species) were higher in the MCII-group. More specifically, these five taxa increases in relative abundance more highly and/or frequently in the MCII-group than in the MCII+ group.

**Figure 4.**
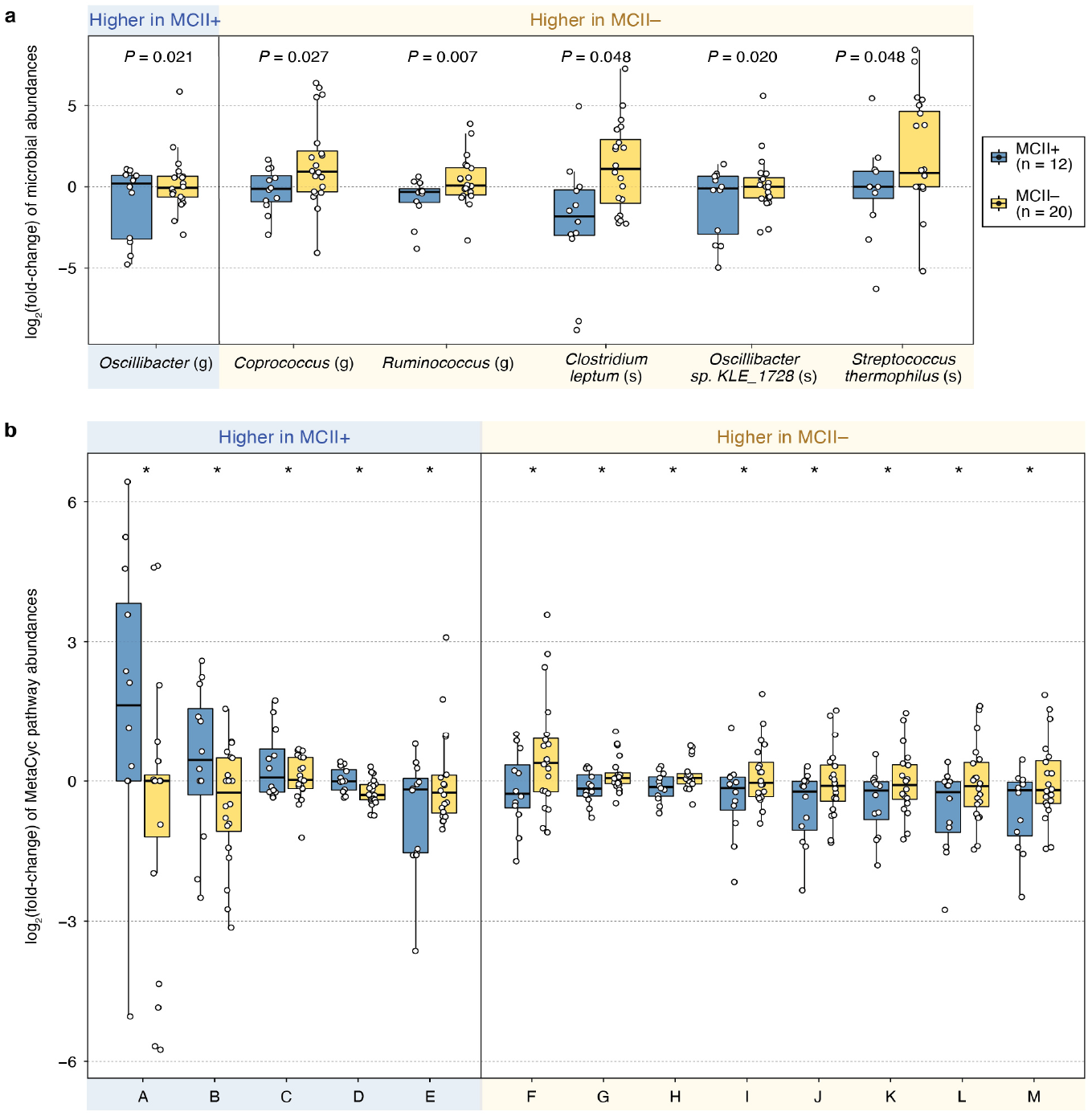
MCII+ and MCII-groups display significantly different fold-changes in microbial taxa and biochemical pathways from baseline to follow-up. **(a)** Six microbial taxa showed significant differences in fold-changes (from baseline to follow-up) between the MCII+ and MCII-patient groups (*P* < 0.05, MLRM). Relative abundances diverged in opposite directions in four taxa: *Oscillibacter* increased (decreased) in relative abundance from baseline to follow-up visit in the MCII+ (MCII-) patient group, while the relative abundances of *Coprococcus, Ruminococcus*, and *Clostridium leptum* decreased (increased) at the follow-up visit in patients of the MCII+ (MCII-) group. **(b)** Thirteen MetaCyc biochemical pathways were identified as having significantly different fold-changes between the two patient groups (*P* < 0.05, MLRM). Pathway B increased (decreased) in relative abundance from baseline to follow-up in the MCII+ (MCII-) group; in contrast, the relative abundance of pathways F, G, and H decreased (increased) at the follow-up visit in the MCII+ (MCII-) group. *P*-values shown above the box plots were found using multiple linear regression models (MLRMs) designed to test for the statistical significance of the relationship between MCII patient group and fold-change in relative abundances of microbial taxa/pathways. These models were controlled for the following patient factors: age group, sex, smoking status, duration (days) between baseline and follow-up visits, baseline CDAI, and use of csDMARDs. ^*^, 0.01 ≤ *P* < 0.05. Taxonomic ranks: c, class; o, order; f, family; g, genus; s, species. MetaCyc pathways: A, Superpathway of Fucose and Rhamnose Degradation; B, ADP-L-glycero- & beta-D-manno-heptose Biosynthesis; C, Pyridoxal 5’-phosphate Biosynthesis I; D, GDP-mannose Biosynthesis; E, Seleno-amino Acid Biosynthesis; F, myo-, chiro- and scillo-inositol Degradation; G, Chorismate Biosynthesis from 3-dehydroquinate; H, Superpathway of Aromatic Amino Acid Biosynthesis; I, Tetrapyrrole Biosynthesis I; J, L-homoserine and L-methionine Biosynthesis; K, Superpathway of L-methionine Biosynthesis; L, L-methionine Biosynthesis I; M, Superpathway of S-adenosyl-L-methionine Biosynthesis.

In the MCII+ group, the relative abundances of four taxa (*Coprococcus, Ruminococcus, Clostridium leptum, Oscillibacter* sp. KLE_1728) decreased from baseline to follow-up (median log_2_(fold-change) ≤ −0.1), whereas *Oscillibacter* increased in abundance (median log_2_(fold-change) ≥ 0.1) (**Fig. 4a, Fig. S4a**). In the MCII-group, the relative abundances of four taxa (*Coprococcus, Ruminococcus, Clostridium leptum, Streptococcus thermophilus*) increased from baseline to follow-up (median log_2_(fold-change) ≥ 0.1), while *Oscillibacter* decreased in abundance (median log_2_(fold-change) ≤ −0.1). Strikingly, these observations imply that the changes in relative abundances of *Coprococcus, Oscillibacter, Ruminococcus*, and *Clostridium leptum* (from baseline to follow-up) in the MCII+ group and those in the MCII-group generally diverged in opposite directions.

Next, we identified thirteen biochemical pathways having significantly different fold-changes between the two MCII patient groups (*P* < 0.05, MLRM; **Fig. 4b, Fig. S4b**): i) five pathways, including those involving sugar metabolism (e.g., fucose and rhamnose degradation, heptose derivative biosynthesis, GDP-mannose biosynthesis), had higher fold-changes in the MCII+ group; and ii) eight pathways, the majority of which for amino acid metabolism (e.g., aromatic amino acid biosynthesis, L-homoserine and L-methionine biosynthesis), had higher fold-changes in the MCII-group.

As seen for microbial taxa, changes in relative abundance of four of these thirteen biochemical pathways were in opposite directions in the two patient groups: ADP-L-glycero- & beta-D-manno-heptose Biosynthesis (**Fig. 4b**, Pathway B) generally increased and decreased in the MCII+ and MCII-group, respectively; myo-, chiro- and scyllo-inositol Degradation (**Fig. 4b**, Pathway F), Chorismate Biosynthesis from 3-dehydroquinate (**Fig. 4b**, Pathway G), and Superpathway of Aromatic Amino Acid Biosynthesis (**Fig. 4b**, Pathway H) generally decreased and increased in the MCII+ and MCII-group, respectively. Although it is yet uncertain why the relative abundances of these particular microbial taxa and biochemical pathways increase (or decrease) in one patient group but decrease (or increase) in the other, such analyses into the *changes* of distinct gut microbiome features, and how these changes are relevant to clinical improvement, can reveal important additional insights not provided by cross-sectional datasets.

**Figure S4.**
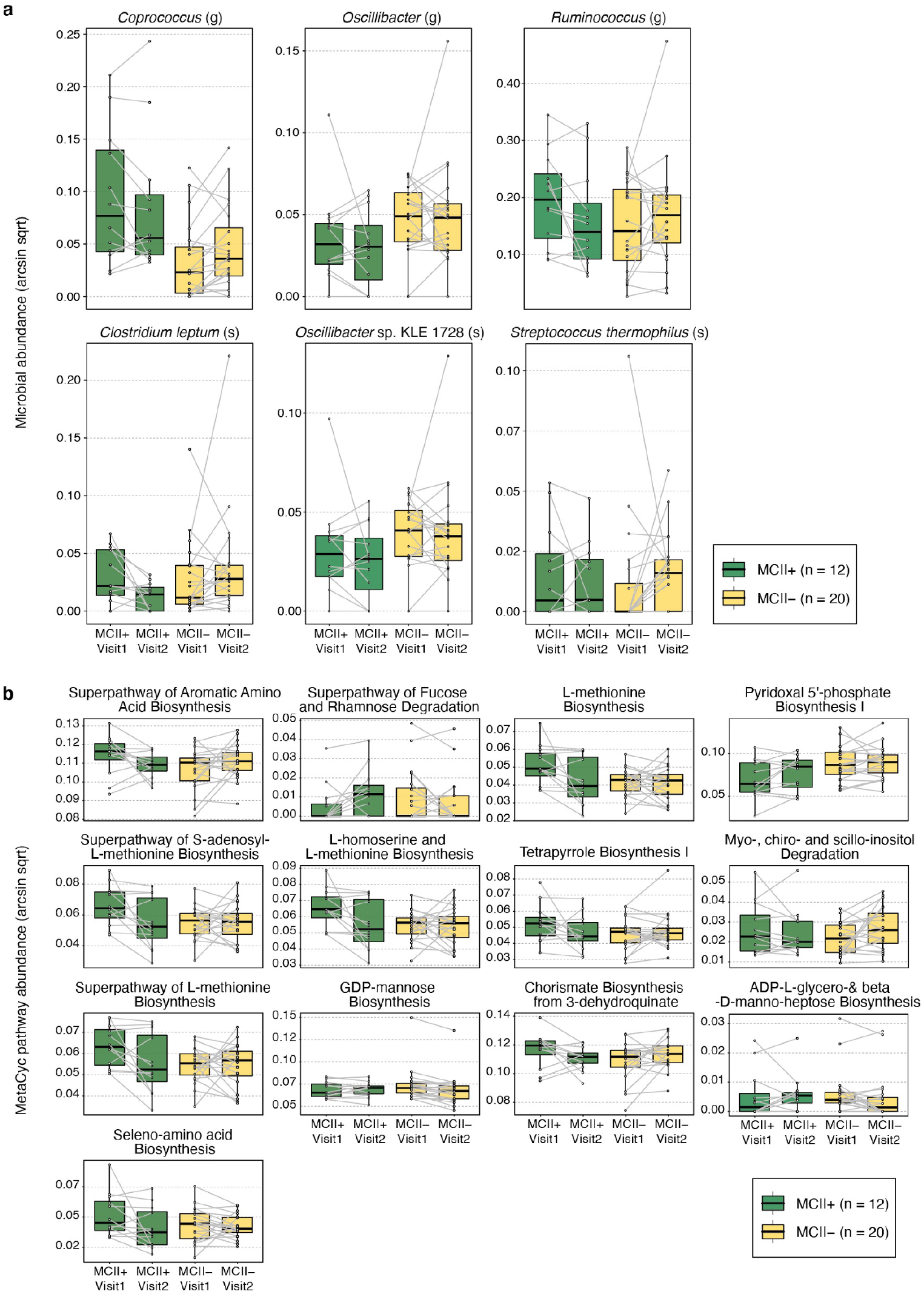
Microbial taxa and biochemical pathways whose change in relative abundance from baseline to follow-up vary differently between MCII patient groups. **(a)** Six microbial taxa (*Coprococcus, Oscillibacter, Ruminococcus, Streptococcus thermophilus, Clostridium leptum* and *Oscillibacter* sp. KLE 1728) displayed significantly different fold-changes between the two MCII patient groups. **(b)** Thirteen MetaCyc biochemical pathways (Superpathway of Aromatic Amino Acid Biosynthesis, Superpathway of Fucose and Rhamnose Degradation, L-methionine Biosynthesis I, Pyridoxal 5’-phosphate Biosynthesis I, Superpathway of S-adenosyl-L-methionine Biosynthesis, L-homoserine and L-methionine Biosynthesis, Tetrapyrrole Biosynthesis I, Myo-, chiro- and scillo-inositol Degradation, Superpathway of L-methionine Biosynthesis, GDP-mannose Biosynthesis, Chorismate Biosynthesis from 3-dehydroquinate, ADP-L-glycero-&beta-D-manno-heptose Biosynthesis and Seleno-amino Acid Biosynthesis) showed significantly different fold-changes between the two patient groups. Points connected by gray lines indicate stool metagenome (gut microbiome) samples from the same patient at two clinical visits. MCII+, patients who showed MCII. MCII-, patients who did not show MCII. Visit 1: baseline; Visit 2: follow-up. Taxonomic ranks: c, class; o, order; f, family; g, genus; s, species.

### Gut microbiome is a predictive marker for clinical improvement and clinical disease activity in patients with RA

Having the capability to reliably predict whether a patient will show clinical improvement—independent of prior treatment and clinical course—would address what has been a steep challenge in the clinical practice of RA. As described above, we identified differences in baseline gut microbiome properties between MCII+ and MCII-patient groups. As an extension of these findings, we next turned to the question of how accurately baseline gut microbiome profiles and clinical and demographic data, combined with a machine-learning approach, can predict MCII class for a particular patient or group of patients; this essentially enables us to forecast whether a patient will have a good prognosis, that is, achieving MCII or not. To this end, we used a neural network classification model that incorporates baseline microbiome, clinical, and demographic data as the input variables to classify patients into one of the two MCII patient groups (**Fig. 5a**; **Methods**). The neural network model was able to distinguish the two groups with reasonably high prediction accuracy in leave-one-out cross-validation: a balanced accuracy (i.e., average of the proportions of MCII+ and MCII-samples that were correctly classified) of 90.0%, as the classification accuracy for the MCII+ and MCII-group was 100.0% (12 of 12) and 80.0% (16 of 20), respectively (**Fig. 5b**). Encouragingly, we were able to correctly predict MCII in all twelve patients who did indeed show clinical improvement.

**Figure 5.**
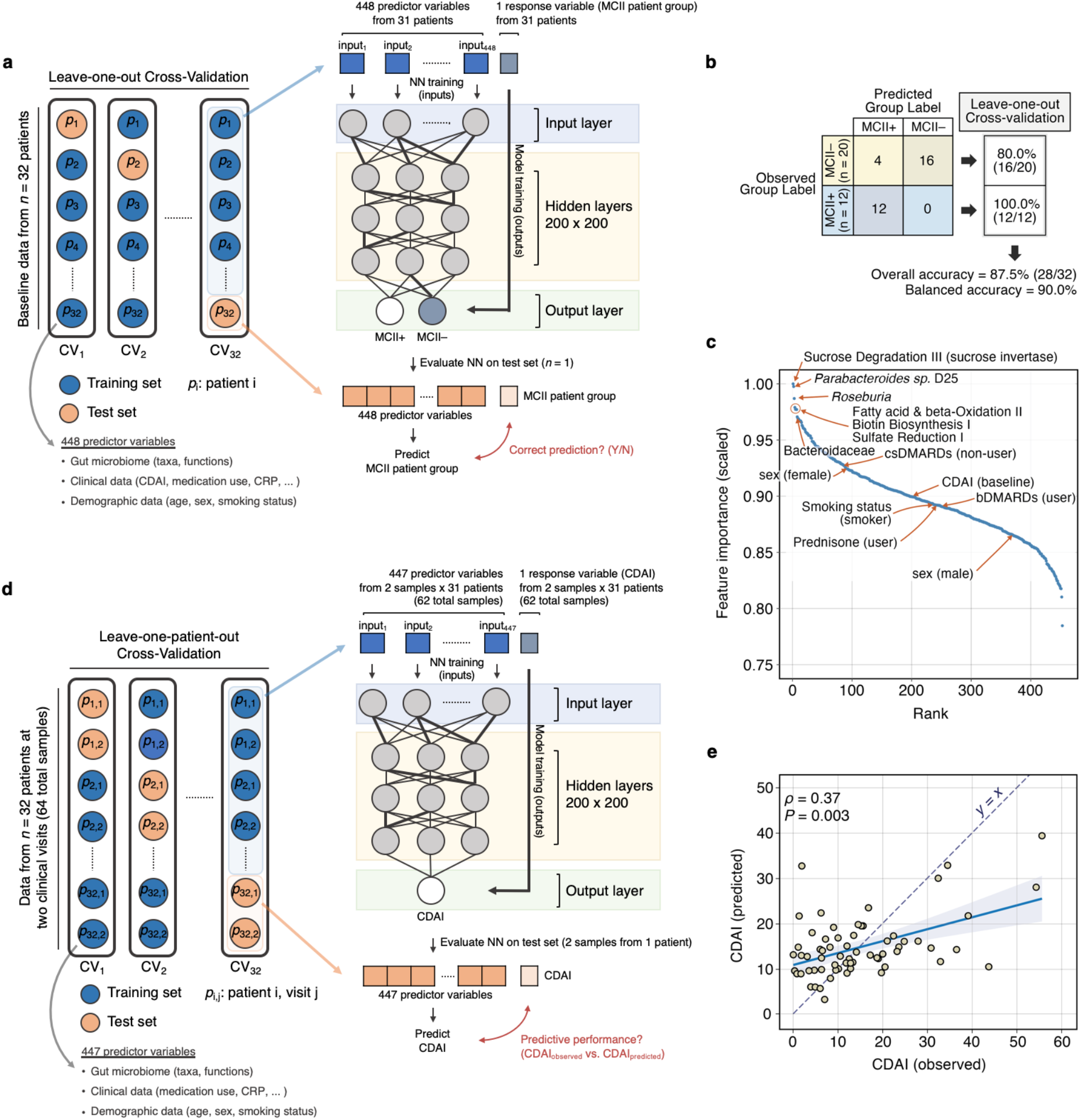
Performance evaluation of neural network-based prediction models in determining minimum clinically important improvement and disease activity score (CDAI). **(a)** A neural network model was designed to classify patients into one of two MCII patient groups using baseline gut microbiome, clinical, and demographic input features. In leave-one-out cross-validation, this resulted in **(b)** a confusion matrix of model predictions showing an overall classification accuracy of 87.5% and a balanced accuracy of 90.0%. MCII+, patients who showed MCII. MCII-, patients who did not show MCII. **(c)** A ranked-order of model input features (total: 448) based upon their scaled (from 0 to 1) importance showing that microbiome data were much more influential contributors to the neural network’s decision-making process than clinical and demographic information. Far left: ranked most important; far right: ranked least important. **(d)** Another neural network model was constructed to predict CDAI from the same input variables (excluding CDAI) in leave-one-patient-out cross-validation. **(e)** CDAI predictions were made on both samples from the same left-out patient in each cross-validation loop (see **Methods**). In the scatter-plot, predictions made across all 32 iterations of cross-validation are shown simultaneously. Overall correlation between observed and predicted scores: Spearman’s *ρ* = 0.37 (*P* = 0.003; 95% confidence interval: [0.12, 0.58]). Dashed violet line indicates ‘y = x’, i.e., an exact match between the observed and predicted values.

Next, by finding which input features were the most informative in the classification process, we rank-ordered all features based upon their scaled importance as determined by the neural network. We found that the top-ranked features were mainly composed of taxonomic and functional components from gut microbiome data (**Fig. 5c**). Of note, the top five important features were the Sucrose Degradation III pathway, *Parabacteroides* sp. D25 (species), *Roseburia* (genus), Fatty Acid & beta-oxidation II pathway, and Biotin Biosynthesis I pathway. Surprisingly, data from clinical and demographic characteristics were ranked much lower; the highest ranked non-microbiome feature was related to the use of csDMARDs, which was ranked 78th (out of 448) in regards to feature importance, followed by sex (female), which was ranked 87th.

Having shown that gut microbiome data can be used to predict whether (or not) a patient will show MCII, we developed another neural network model to evaluate how well the aforementioned predictor variables can predict CDAI (**Fig. 5d**; **Methods**). The direct prediction of a clinical disease activity score using gut microbiome has yet to be performed in any chronic disease, although a previous study by Tedjo *et al*. used a Random Forests classifier with operational taxonomic units (OTUs) of the gut microbiome in Crohn’s Disease to differentiate between active disease and remission [56]. By using a leave-one-patient-out cross-validation scheme, wherein predictions in each cross-validation loop were made on both samples from a single left-out patient (**Methods**), we found that our neural network achieved a moderate, yet significant, correlation between observed (actual) and predicted CDAI (Spearman’s *ρ* = 0.37, *P* = 0.003; **Fig. 5e**). Interestingly, the predicted CDAI fits a lower slope compared to the slope of an exact match between observed and predicted values. CDAI beyond ∼15 were under-predicted, whereas CDAI below ∼15 were over-predicted; this threshold could possibly indicate a breakpoint at which our model exhibits different relationships between the response and predictor variables. In summary, the gut microbiome shows promise as a non-invasive screening tool for predicting clinical improvement and perhaps also for monitoring RA disease activity.

## Discussion

To the best of our knowledge, this is the first study to date that uses shotgun metagenomic sequencing of stool to investigate the ties between the gut microbiome and MCII in RA disease activity. This study addresses the following key questions: What are the distinct microbes and functions that define gut ecologies in patients who achieve MCII compared to patients who do not? Are these specific gut microbiome “signatures” predictive of MCII? Or in other words, how well does the gut microbiome forecast the trajectory of RA disease activity irrespective of prior clinical course? To this end, we compared the baseline gut microbiome compositions between RA patients who eventually showed improvement in disease activity and those who did not. First, we found that the status of MCII is a significant contributor to the variation in gut microbiome community composition. Next, a more detailed examination of baseline gut microbiomes allowed us to identify higher species-level alpha-diversity and beta-diversity in the MCII+ group (i.e., patients who showed MCII) than in the MCII-group (i.e., patients who did not show MCII). Additionally, we identified several microbial taxa and microbiome-derived MetaCyc biochemical pathways as differentially abundant between the two MCII patient groups. Furthermore, we observed several taxa and pathways as having significant differences in fold-change (from baseline to follow-up) between the two patient groups. Lastly, we demonstrate that the integration of gut microbiome and machine-learning technology could theoretically be an avenue for the prediction of disease course in RA. More specifically, by incorporating baseline microbiome, clinical, and demographic data into a deep-learning neural network, we were able to effectively classify patients into their MCII+ or MCII-group, thereby allowing us to forecast MCII in patients with RA. With further development, such prognostic biomarkers could identify patients who will achieve MCII earlier on and spare them the expense and risk of aggressive therapies; conversely, such tools can detect patients whose disease symptoms are less likely to improve, and perhaps allow clinicians to target and monitor them more closely. In all, our proof-of-concept study targets a significant unmet medical need in RA, and demonstrates the utility of the gut microbiome for the precision medicine era.

We identified several microbial taxa at baseline, including *Coprococcus, Bilophila* sp. 4_1_30, and Prevotellaceae, to have significantly different relative abundances between the MCII+ and MCII-patient groups. *Coprococcus* was found to be relatively higher in the MCII+ group compared to the MCII-group. Microorganisms of this genus are known to produce butyrate, which is known for its anti-inflammatory effects [57–63]. For example, a study in mice showed that butyrate can suppress inflammation by inhibiting histone deacetylases (HDACs) in bone marrow cells [58]. Previously, the administration of an HDAC inhibitor *in vivo* was found to promote the production and suppressive function of Foxp3^+^ regulatory T (T_reg_) cells [64]. The anti-inflammatory effect of butyrate was also shown in *Staphylococcus aureus* cell-stimulated human monocytes, to which adding butyrate led to a reduction and increase of proinflammatory cytokine IL-12 and anti-inflammatory cytokine IL-10, respectively [59]. In addition, *Bilophila* sp. 4_1_30 was found to be higher in patients of the MCII+ group. The role of *Bilophila* species in inflammatory or auto-immune diseases is not yet fully understood. A couple of studies have shown the positive association of *Bilophila* species (in particular *B. wadsworthia*) with pro-inflammatory immune responses [65,66], while another study has shown that *Bilophila* species have negative associations with LPS-induced, TNFα-mediated immune responses in whole blood peripheral blood mononuclear cells [67]. Lastly, Prevotellaceae was also found to have greater abundance in the MCII+ group. Some species in this family are known for their pro-inflammatory effects [68,69]; therefore, this observation possibly suggests that host immune responses to Prevotellaceae are specific to particular species and/or strains [70].

In addition to baseline differences in microbial taxa between the MCII+ and MCII-groups, we observed differences in the abundances of thirteen biochemical pathways at baseline. Eight of these differentially-abundant pathways are involved in the biosynthesis of amino acids, such as arginine, methionine, ornithine, and tryptophan. All two pathways involved in methionine biosynthesis were found to be more abundant in the MCII+ group. Interestingly, dietary supplementation with high levels of methionine has been shown to attenuate arthritis severity in arthritic rats, and also to increase levels of serum Insulin-like Growth Factor-1 (IGF-I) [71]; and to this point, IGF-I was previously found to be significantly lower in female patients with RA than in controls [72]. Alternatively, all four arginine biosynthesis pathways were of lower abundance in the MCII+ group. A recently published study has shown that restriction of arginine improves outcome in multiple murine arthritis models by controlling the metabolism and formation of multi-nuclear giant cells [73]. Collectively, our results implicate various aspects of the gut microbiome with improvement in chronic, debilitating symptoms in RA, raising the interesting possibility of intervening on these markers, e.g., introducing specific desirable bacterial strains into the gut or targeting certain microbial metabolic pathways as a basis for drug development.

Several limitations should be acknowledged when interpreting our results. First and foremost, the relatively small sample size used in our study limits generalization of the findings to the broader range of RA conditions. It was beyond the scope of this observational cohort study to restrict the time of follow-up between clinical visits, leading to variability in the duration of follow-up. While this study is the first to associate gut microbiome signatures with MCII in RA, we do note that our results were derived from a pilot cohort of 32 patients; therefore, conducting more analyses and validation on larger cohorts with pre-specified clinical endpoints is the crucial next step to strengthen and confirm our findings. Second, our results could be influenced by confounders inherent to our cohort of patients. We do acknowledge that there may be geographical/cultural biases in our results, since the patients included in this study are mostly from the midwest region of the United States. Our statistical methods to identify associations between the gut microbiome and MCII were controlled for age, sex, smoking status, baseline disease activity, follow-up duration, and medication use. However, dietary habits were not assessed, which is a variable well known to influence the composition of the gut microbiome [74,75]. Importantly, we were not able to statistically control for patient BMI, as current height and weight were found to be missing in several patient records. Of note, obesity is not only strongly tied to gut microbiome [76–78], but also known as a prognostic factor in RA. More specifically, patients with obesity have been found to be less likely to respond to disease-modifying therapy [79]. How much BMI plays a role in shaping the current results will be addressed in our future studies. Third, as is often the case in retrospective cohort studies, we cannot completely eliminate the possibility of patient selection bias. For example, patients may not elect to return for a follow-up visit depending on a certain disease severity. Additionally, among the patients whose clinical samples were available in our biobank, some clinical/demographic data were incomplete for both time-points. Such reasons result in exclusion of these patients from our study, and therefore may bias the type of patients who were analyzed. Fourth, all descriptions of annotated biochemical pathways of the gut microbiome allude to *functional potential*, that is, functional possibilities derived from genetic content. We did not employ transcriptomics or proteomics technologies to assess enzyme abundances; metabolomics to detect small-molecules; or cellular assays to determine metabolic flux. However, these are all promising methods that we can later use to obtain much richer insight into how microbial metabolism affects RA disease course. Fifth, clearly our study cannot provide causal mechanisms underlying the associations between the gut microbiome and MCII in RA disease activity. However, a closer investigation on particular microbial taxa or microbiome-derived pathways identified in our study may provide a promising launchpad for future studies delving into specifically how alterations in the gut microbiome influence RA-associated changes in human physiology or in systemic, chronic inflammation. Sixth, all predictions regarding MCII patient group and CDAI were performed in cross-validation on the original discovery cohort. It remains to be seen how well the robustness of our prediction models will hold up when demonstrated on an independent validation cohort once available. Finally, although we found that the gut microbiome is surprisingly predictive of MCII, our study is limited by the fact that we collected stool samples and assessed patients’ disease activity at only two time-points. It could be possible that associations between gut microbiome and MCII may not persist past the second visit. Surely, future studies extending this current work will need to entail having larger cohorts, patients with new onset RA, and several longitudinal sample collections, while considering more potentially confounding factors (e.g., geography, race/ethnicity, diet, and lifestyle).

## Conclusions

Several aspects of the gut microbiome are associated with future prognosis in RA, providing motivation for further studies on the effect of intestinal microflora and various patient factors on autoimmune response and clinical course. Additionally, shotgun metagenomic sequencing of microbial communities in stool samples can serve as an effective and reliable predictor of whether patients with RA will achieve clinically important improvement in disease activity. Therefore, learning to better “read” the gut microbiome and its changes, as well as mapping its complex relationship with disease symptoms, may provide a promising route for making more accurate clinically-informed decisions for RA patients. Ultimately, we expect our work to be one cornerstone for a suite of new, omics-based clinical tools to aid in early detection, diagnosis, prognosis, and treatment in RA. Looking ahead, possible solutions to treat chronic auto-immune or inflammatory diseases could well involve modifying the gut microbiome to an ecological state primed to enhance clinical outcome.

## Data Availability

Sequencing data for stool metagenomes used in this study have been deposited at NCBI Sequence Read Archive (SRA) data repository (BioProject number PRJNA598446 and PRJNA687957) and can be downloaded without any restrictions. The deposited sequences include .fastq files for 64 stool metagenomes collected from 32 patients with rheumatoid arthritis. Human reads were identified and removed prior to data upload.

## Availability of data and materials

Sequencing data for stool metagenomes used in this study have been deposited at NCBI’s Sequence Read Archive (SRA) data repository (BioProject number PRJNA598446 and PRJNA687957), and can be downloaded without any restrictions. The deposited sequences include .fastq files for 64 stool metagenomes collected from 32 patients with rheumatoid arthritis. Human reads were identified and removed prior to data upload.

## Competing interests

J.M.D. has a research grant from Pfizer. All other authors declare that they have no competing interests.

## Funding

This work was supported in part by the Mayo Clinic Center for Individualized Medicine (to V.K.G., K.C., U.B., B.H., and J.S.), and Mark E. and Mary A. Davis to Mayo Clinic Center for Individualized Medicine (J.M.D., J.S.).

## Authors’ contributions

V.K.G., J.M.D., and J.S. conceived the problem. V.K.G., K.Y.C., J.M.D., and J.S. designed all analytical methodologies. V.K.G., K.Y.C., and U.B. performed the computational analyses. All authors analyzed and reviewed the data, and provided critical feedback. V.K.G., K.Y.C, J.M.D., and J.S. wrote the manuscript, with editorial contributions from other authors. J.M.D. is the principal investigator of the Mayo Clinic Rheumatology Biobank, from which stool samples were collected from patients with rheumatoid arthritis. All authors reviewed and approved the final manuscript.

## Acknowledgments

First and foremost, we thank our dear patients who volunteered for this study. We also thank the Mayo Clinic Division of Rheumatology study coordinators (Jennifer Sletten and Kathleen McCarthy-Fruin) and the Mayo Clinic Medical Genome Facility staff members (Julie Lau, Jeffrey Meyer, and Bruce Eckloff) for making this work possible.

## Notes

### Author Declarations

The study was approved by the Mayo Clinic Institutional Review Board (no. 14-000616).

